# The Humoral Response to the BNT162b2 Vaccine in Hemodialysis Patients

**DOI:** 10.1101/2021.05.24.21257425

**Authors:** Kevin Yau, Kento T. Abe, David Naimark, Matthew J. Oliver, Jeffrey Perl, Jerome A. Leis, Shelly Bolotin, Vanessa Tran, Sarah Mullin, Ellen Shadowitz, Julie Garnham-Takaoka, Keelia Quinn de Launay, Alyson Takaoka, Sharon E. Straus, Allison J. McGeer, Christopher T. Chan, Karen Colwill, Anne-Claude Gingras, Michelle A. Hladunewich

## Abstract

**Importance:** Hemodialysis patients have an exceptionally high mortality from COVID-19 and this patient population often has a poor response to vaccinations. Randomized controlled trials for COVID-19 vaccines included few patients with kidney disease, therefore vaccine immunogenicity is uncertain in this population.

**Objective:** Evaluate the SARS-CoV-2 antibody response in chronic hemodialysis patients following one versus two doses of BNT162b2 COVID-19 vaccination compared to health care worker controls and convalescent serum.

**Design:** Prospective observational cohort study.

**Setting:** Single centre study in Toronto, Ontario, Canada.

**Participants:** 142 in-centre hemodialysis patients and 35 health care worker controls.

**Exposure:** BNT162b2 (Pfizer-BioNTech) COVID-19 vaccine.

**Main Outcomes and Measures:** SARS-CoV-2 IgG antibodies to the spike protein (anti-spike), receptor binding domain (anti-RBD), and nucleocapsid protein (anti-NP) were measured in 66 hemodialysis patients receiving one vaccine dose following a public health policy change, 76 patients receiving two vaccine doses, and 35 health care workers receiving two vaccine doses.

**Results:** Detectable anti-NP suggestive of natural SARS-CoV-2 infection was detected in 15/142 (11%) of patients at baseline while only three patients had prior RT-PCR confirmed COVID-19. Two additional patients contracted COVID-19 after receiving two doses of vaccine. In patients receiving a single BNT162b2 dose, seroconversion occurred in 53/66 (80%) for anti-spike and 35/66 (55%) for anti-RBD by 28 days post dose, but only 15/66 (23%) and 4/66 (6%), respectively attained a robust response as defined by reaching the median level of anti-spike and anti-RBD in convalescent serum from COVID-19 survivors. In patients receiving two doses of BNT162b2 vaccine, seroconversion occurred in 69/72 (96%) for anti-spike and 63/72 (88%) for anti-RBD by 2 weeks following the second dose while 52/72 (72%) and 43/76 (41%) reached the median convalescent serum level of anti-spike and anti-RBD, respectively. In contrast, 35/35 (100%) of health care workers exceeded the median level of anti-spike and anti-RBD found in convalescent serum 2-4 weeks after the second dose.

**Conclusions and Relevance:** This study confirms poor immunogenicity 28 days following a single dose of BNT162b2 vaccine in the hemodialysis population, supporting adherence to recommended vaccination schedules, and avoiding delay of the second dose in these at-risk individuals.

**Key Points:** *Question:* What is the serologic response to the BNT162b2 COVID-19 vaccine in hemodialysis patients?

*Findings:* In this prospective observational study, humoral response was compared in 66 hemodialysis patients sampled 28 days after receipt of one dose of vaccine to 76 patients who received two doses of vaccine sampled 14 days after the second dose. Among those receiving one dose, 6% had anti-RBD response above the median level of convalescent serum versus 41% who received two doses.

*Meaning:* Given that hemodialysis patients exhibit a poor humoral response to a single dose of BNT162b2 vaccine, the second dose should not be delayed.

## Introduction

Severe acute respiratory syndrome coronavirus 2 (SARS-CoV-2), which causes coronavirus disease 2019 (COVID-19), has resulted in a devastating global pandemic. Among those most severely affected are patients on maintenance hemodialysis who must visit facilities at least thrice weekly for life sustaining treatment resulting in a five times greater risk for infection than the general population.^1^ Despite adherence to public health guidance, outbreaks have occurred in dialysis units.^2^ Further, patients on dialysis are at greater risk for severe COVID-19, with 63% of chronic dialysis patients with COVID-19 requiring hospitalization and a case fatality rate of 29% in Ontario, Canada.^1^ Confirmatory data from the US Renal Data System found mortality among hemodialysis patients in early 2020 was 16-37% higher than in 2017-2019.^3^

Dialysis patients frequently have diminished immune response to vaccination compared with the general population, as observed during hepatitis B vaccination.^4^ Studies of natural COVID-19 infection in dialysis patients found waning antibody concentrations by three months, raising the possibility that dialysis patients may not develop an adequate vaccination response.^5^ Finally, randomized controlled trials for the BNT162b2 vaccine included few patients with kidney disease.^6^ Therefore data on vaccine immunogenicity are lacking in this high-risk population.

Two doses of BNT162b2 vaccine were administered 21 days apart in randomized controlled trials. However, due to vaccine shortages, countries including the United Kingdom and Canada have prioritized first dose vaccination of the general population,^7^ while delaying the second dose for up to 3-4 months, offering a natural experiment for comparison of one versus two doses. To investigate the humoral response conferred by COVID-19 vaccination in the hemodialysis population, we conducted a prospective observational study measuring SARS-CoV-2 IgG antibody levels following one versus two doses of vaccine.

### Study Design and Methods

In-centre hemodialysis patients age ≥18 years, including those with prior COVID-19, were eligible for this single centre prospective observational study to evaluate SARS-CoV-2 antibody response to the BNT162b2 COVID-19 vaccine (Pfizer-BioNTech). Recruitment of 142 participants occurred between February 2, 2021 and March 3, 2021 at Sunnybrook Health Sciences Centre. A subset of patients (n=76) received two dose vaccination with the second dose a mean of 21 days (range 19-28) following first dose, while 66 patients received a single vaccine dose due to a policy change. In the two-dose group, baseline was prior to second dose and antibody levels were collected weekly until 14 days post second vaccine dose. In those receiving a single vaccine dose, antibody levels were measured at baseline prior to vaccination and 28 days following the first dose. A written questionnaire captured vaccination-related adverse events. Health care worker (HCW) controls received two doses of BNT162b2 vaccine with antibodies measured 2-4 weeks following second dose. This study was approved by the Sunnybrook Health Sciences and Mount Sinai Hospital Research Ethics Board. Written informed consent was obtained from all participants.

Antibodies targeting the full-length spike protein (anti-spike) and its Receptor Binding Domain (anti-RBD) measured humoral response to SARS-CoV-2 vaccination and/or natural infection while antibodies to the nucleocapsid protein (anti-NP) detected natural SARS-CoV-2 infection, as this antigen is not targeted by the BNT162b2 vaccine. SARS-CoV-2 IgG antibodies were measured on a custom automated enzyme-linked immunosorbent assay (ELISA) platform; the sensitivity and specificity of each assay was determined by precision-recall analysis from pre-COVID negative and convalescent controls.^8,9^ Antibody levels were reported as relative ratios to a synthetic standard included as a calibration curve on each assay plate. Thresholds for positivity (seroconversion) were determined by aggregating data from negative controls and calculating the mean + 3 standard deviations. Relative antibody levels were also compared to the median levels of convalescent serum taken 21-115 days post symptom onset in patients with COVID 19; expression of vaccination-induced antibody levels to convalescent individuals helps to define correlates of protection.^10^ The association between reactogenicity following second vaccine dose and the proportion of patients with anti-spike or anti-RBD seroconversion two weeks following second dose was assessed by a Chi-Squared test.

## Results

Among 142/157 (90%) consenting in-centre hemodialysis patients, the median age was 72 years and 48/142 (34%) were female **(Table 1)**. At baseline, 15/142 (11%) of patients had detectable anti-NP of whom 3/142 (2%) patients had RT-PCR confirmed COVID-19. Characteristics of patients receiving one versus two vaccine doses were similar. HCW controls had a median age of 46, 33/35 (94%) were female, and 3/35 (9%) had prior COVID-19.

**Table 1.**
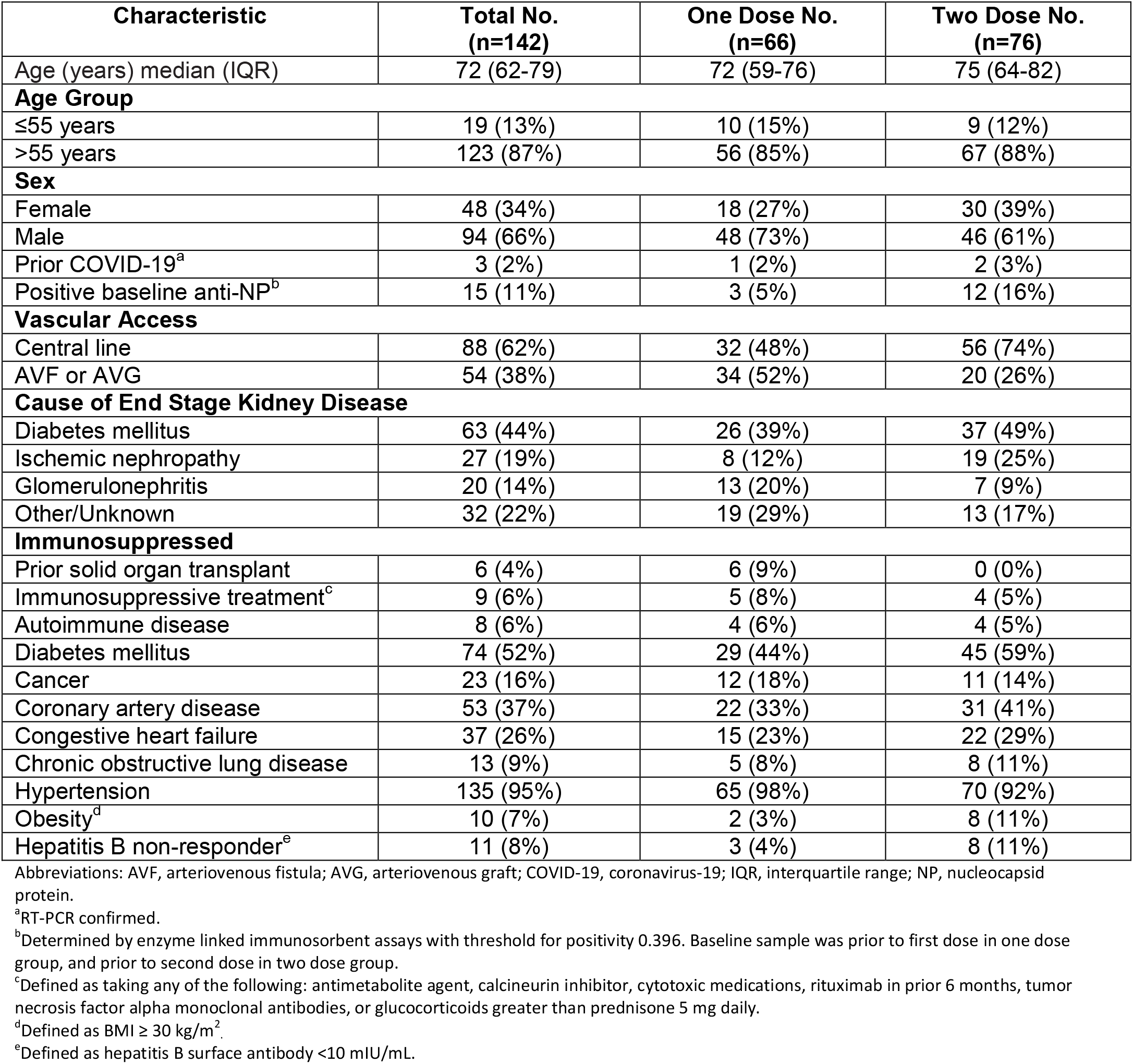
Clinical Characteristics of 142 Hemodialysis Patients Receiving BNT162b2 Vaccine

| Characteristic | Total No.<br>(n=142) | One Dose No.<br>(n=66) | Two Dose No.<br>(n=76) |
| --- | --- | --- | --- |
| Age (years) median (IQR) | 72 (62-79) | 72 (59-76) | 75 (64-82) |
| **Age Group** |  |  |  |
| ≤55 years | 19 (13%) | 10 (15%) | 9 (12%) |
| >55 years | 123 (87%) | 56 (85%) | 67 (88%) |
| **Sex** |  |  |  |
| Female | 48 (34%) | 18 (27%) | 30 (39%) |
| Male | 94 (66%) | 48 (73%) | 46 (61%) |
| Prior COVID-19<sup>a</sup> | 3 (2%) | 1 (2%) | 2 (3%) |
| Positive baseline anti-NP<sup>b</sup> | 15 (11%) | 3 (5%) | 12 (16%) |
| **Vascular Access** |  |  |  |
| Central line | 88 (62%) | 32 (48%) | 56 (74%) |
| AVF or AVG | 54 (38%) | 34 (52%) | 20 (26%) |
| **Cause of End Stage Kidney Disease** |  |  |  |
| Diabetes mellitus | 63 (44%) | 26 (39%) | 37 (49%) |
| Ischemic nephropathy | 27 (19%) | 8 (12%) | 19 (25%) |
| Glomerulonephritis | 20 (14%) | 13 (20%) | 7 (9%) |
| Other/Unknown | 32 (22%) | 19 (29%) | 13 (17%) |
| **Immunosuppressed** |  |  |  |
| Prior solid organ transplant | 6 (4%) | 6 (9%) | 0 (0%) |
| Immunosuppressive treatment<sup>c</sup> | 9 (6%) | 5 (8%) | 4 (5%) |
| Autoimmune disease | 8 (6%) | 4 (6%) | 4 (5%) |
| Diabetes mellitus | 74 (52%) | 29 (44%) | 45 (59%) |
| Cancer | 23 (16%) | 12 (18%) | 11 (14%) |
| Coronary artery disease | 53 (37%) | 22 (33%) | 31 (41%) |
| Congestive heart failure | 37 (26%) | 15 (23%) | 22 (29%) |
| Chronic obstructive lung disease | 13 (9%) | 5 (8%) | 8 (11%) |
| Hypertension | 135 (95%) | 65 (98%) | 70 (92%) |
| Obesity<sup>d</sup> | 10 (7%) | 2 (3%) | 8 (11%) |
| Hepatitis B non-responder<sup>e</sup> | 11 (8%) | 3 (4%) | 8 (11%) |

Abbreviations: AVF, arteriovenous fistula; AVG, arteriovenous graft; COVID-19, coronavirus-19; IQR, interquartile range; NP, nucleocapsid protein.
<sup>a</sup>RT-PCR confirmed.
<sup>b</sup>Determined by enzyme linked immunosorbent assays with threshold for positivity 0.396. Baseline sample was prior to first dose in one dose group, and prior to second dose in two dose group.
<sup>c</sup>Defined as taking any of the following: antimetabolite agent, calcineurin inhibitor, cytotoxic medications, rituximab in prior 6 months, tumor necrosis factor alpha monoclonal antibodies, or glucocorticoids greater than prednisone 5 mg daily.
<sup>d</sup>Defined as BMI ≥ 30 kg/m<sup>2</sup>.
<sup>e</sup>Defined as hepatitis B surface antibody <10 mIU/mL.

In 66 patients receiving one vaccine dose, four weeks following the first vaccination, 53/66 (80%) of the patients seroconverted, but only 15/66 (23%) had anti-spike antibodies exceeding the median relative ratio of convalescent individuals (**Table 2)**. In the 76 patients receiving two vaccine doses, the first dose similarly elicited anti-spike seroconversion in 65/76 (86%) with 19/76 (25%) reaching convalescent levels 28 days post dose. The second vaccine dose, however, induced a more robust response increase with 43/76 (57%) reaching convalescent levels by one week further increasing to 52/72 (72%) by 2 weeks with 69/72 (96%) reaching seroconversion **(Figure 1)**.

**Table 2.**
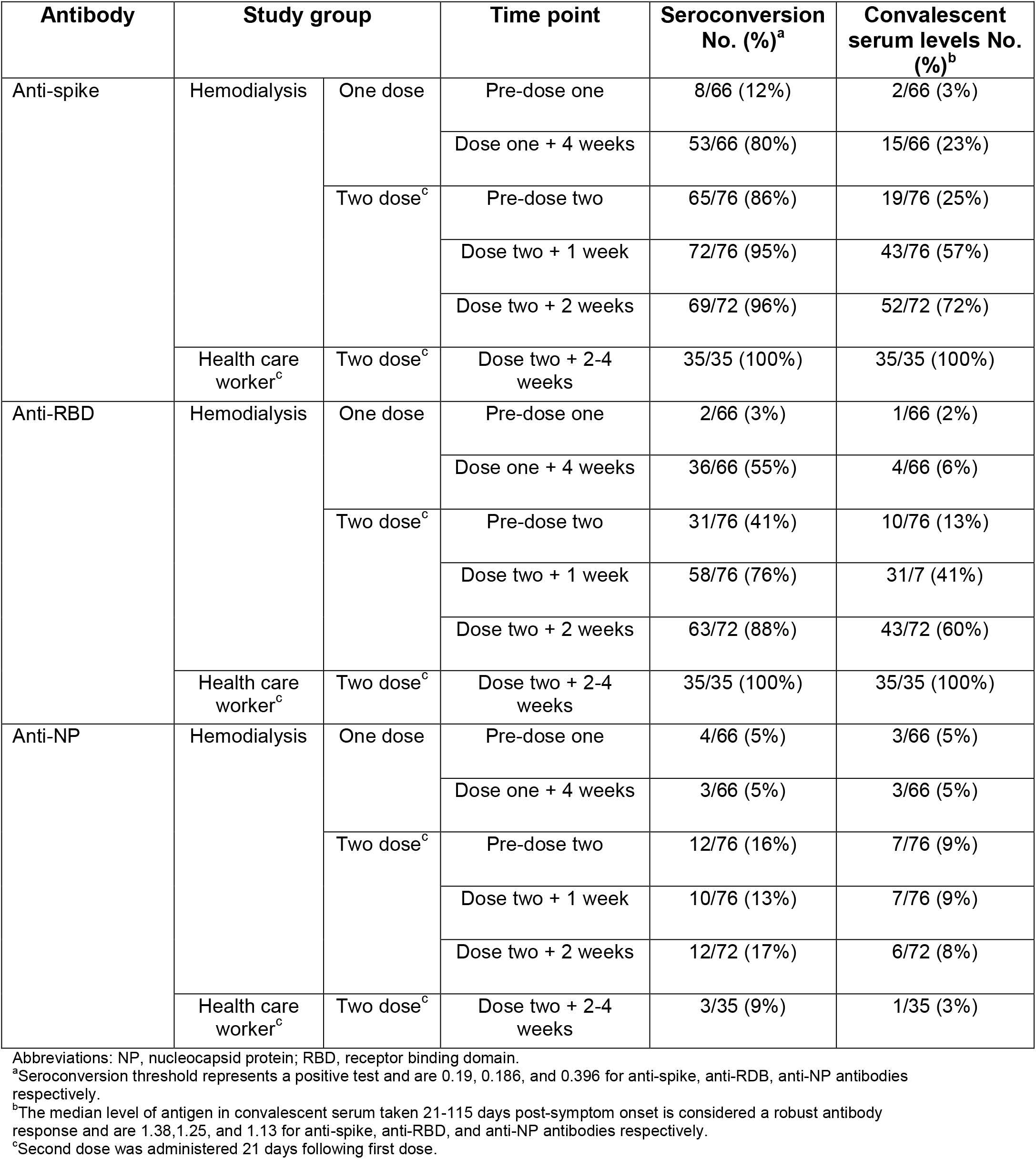
Rates Seroconversion and Attaining Convalescent Serum Levels for SARS-CoV-2 IgG Spike, RBD, and NP Antibodies in Hemodialysis Patients Receiving One Versus Two Doses of BNT162b2

| Antibody | Study group |  | Time point | Seroconversion No. (%)<sup>a</sup> | Convalescent serum levels No. (%)<sup>b</sup> |
| --- | --- | --- | --- | --- | --- |
| Anti-spike | Hemodialysis | One dose | Pre-dose one | 8/66 (12%) | 2/66 (3%) |
|  |  |  | Dose one + 4 weeks | 53/66 (80%) | 15/66 (23%) |
|  |  | Two dose<sup>c</sup> | Pre-dose two | 65/76 (86%) | 19/76 (25%) |
|  |  |  | Dose two + 1 week | 72/76 (95%) | 43/76 (57%) |
|  |  |  | Dose two + 2 weeks | 69/72 (96%) | 52/72 (72%) |
|  | Health care worker<sup>c</sup> | Two dose<sup>c</sup> | Dose two + 2-4 weeks | 35/35 (100%) | 35/35 (100%) |
| Anti-RBD | Hemodialysis | One dose | Pre-dose one | 2/66 (3%) | 1/66 (2%) |
|  |  |  | Dose one + 4 weeks | 36/66 (55%) | 4/66 (6%) |
|  |  | Two dose<sup>c</sup> | Pre-dose two | 31/76 (41%) | 10/76 (13%) |
|  |  |  | Dose two + 1 week | 58/76 (76%) | 31/7 (41%) |
|  |  |  | Dose two + 2 weeks | 63/72 (88%) | 43/72 (60%) |
|  | Health care worker<sup>c</sup> | Two dose<sup>c</sup> | Dose two + 2-4 weeks | 35/35 (100%) | 35/35 (100%) |
| Anti-NP | Hemodialysis | One dose | Pre-dose one | 4/66 (5%) | 3/66 (5%) |
|  |  |  | Dose one + 4 weeks | 3/66 (5%) | 3/66 (5%) |
|  |  | Two dose<sup>c</sup> | Pre-dose two | 12/76 (16%) | 7/76 (9%) |
|  |  |  | Dose two + 1 week | 10/76 (13%) | 7/76 (9%) |
|  |  |  | Dose two + 2 weeks | 12/72 (17%) | 6/72 (8%) |
|  | Health care worker<sup>c</sup> | Two dose<sup>c</sup> | Dose two + 2-4 weeks | 3/35 (9%) | 1/35 (3%) |

Abbreviations: NP, nucleocapsid protein; RBD, receptor binding domain.
<sup>a</sup>Seroconversion threshold represents a positive test and are 0.19, 0.186, and 0.396 for anti-spike, anti-RBD, anti-NP antibodies respectively.
<sup>b</sup>The median level of antigen in convalescent serum taken 21-115 days post-symptom onset is considered a robust antibody response and are 1.38, 1.25, and 1.13 for anti-spike, anti-RBD, and anti-NP antibodies respectively.
<sup>c</sup>Second dose was administered 21 days following first dose.

**Figure 1:**
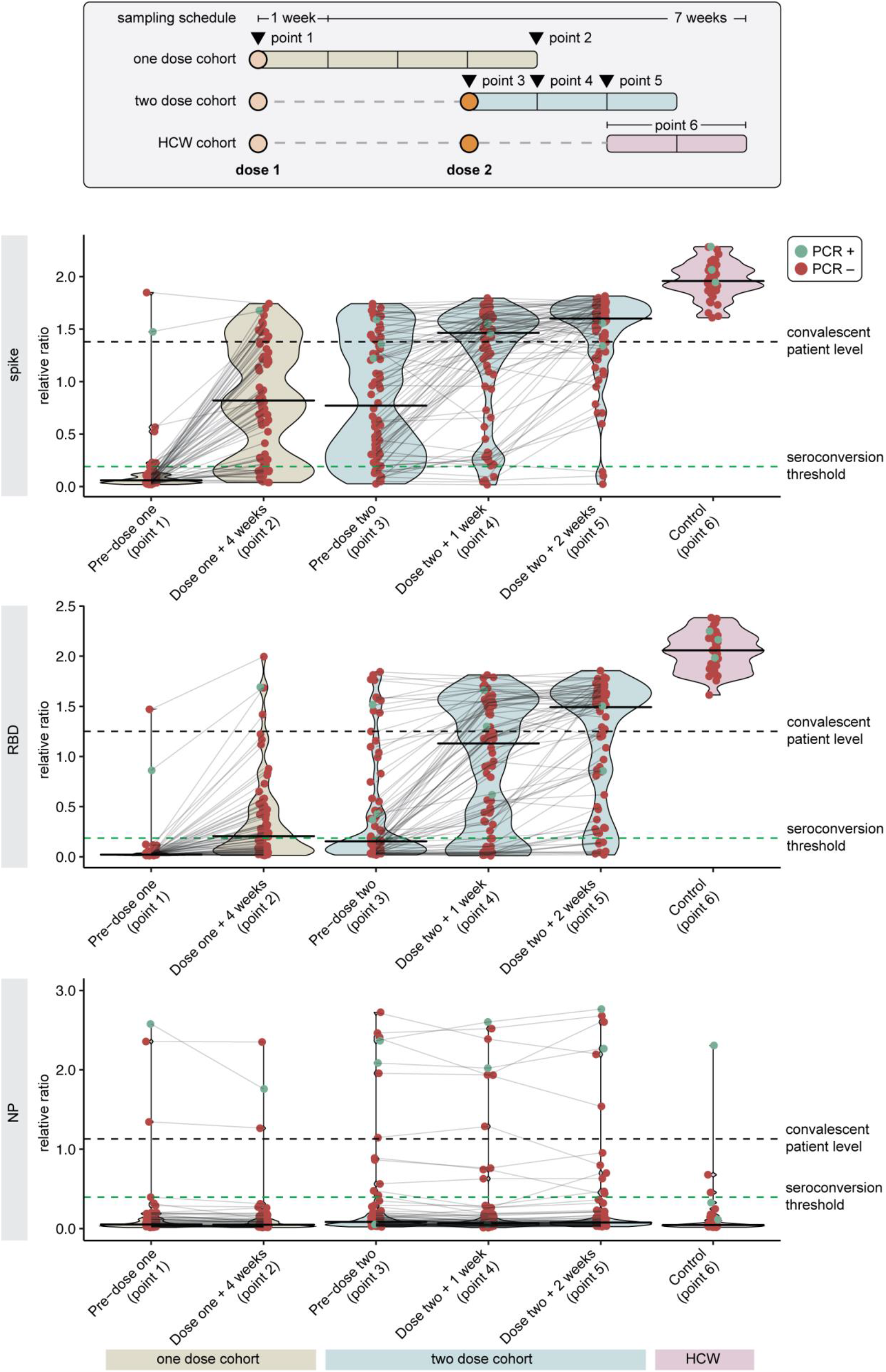
SARS-CoV-2 IgG Spike, RBD, and NP Antibody Response Following One Versus Two Dose BNT162b2 Vaccine in Hemodialysis Patients. Antibody levels are reported as relative ratios to synthetic standards. The samples are grouped into two cohorts who received one (n=66) or two doses (n=76) of vaccine. Dots represent individual serum samples collected at the indicated times, and the samples from the same patients are connected by lines. Green dots indicate individuals with prior RT-PCR confirmed SARS-CoV-2 infection. Seroconversion threshold represents a positive test and are 0.19, 0.186, and 0.396 for anti-spike, anti-RDB, anti-NP antibodies, respectively. The median level of antigen in convalescent serum taken 21-115 days post-symptom onset is considered a robust antibody response and is 1.38, 1.25, and 1.13 for anti-spike, anti-RBD, and anti-NP antibodies, respectively. Healthy control workers (n=35) received two doses of vaccines with serologic measurement 2-4 weeks following dose 2. Abbreviations: HCW, health care worker; NP, nucleocapsid protein; RBD, receptor binding domain.

The same overall trends were more pronounced for antibodies to RBD, which are well correlated with neutralizing antibodies.^9,11^ The first vaccine dose elicited a poor anti-RBD response with seroconversion of 36/66 (55%) in the one dose group and 31/76 (41%) in the two dose group. After one dose, only 4/66 (6%) in the one dose group and 10/76 (13%) in the two-dose group reached convalescent serum levels. However, one week after the second dose of vaccine, 58/76 (76%) of the individuals had seroconverted, with 31/76 (41%) having antibody levels above the median of convalescent serum. Two weeks after second dose of vaccine, 63/72 (88%) seroconverted and 43/72 (60%) were above convalescent serum. In HCW controls, 100% reached the convalescent levels for both anti-spike and anti-RBD 2-4 weeks after two doses. Results were similar when individuals with baseline anti-NP seroconversion were excluded (**eTables 1-2** and **eFigure 1** in the Supplement).

The presence of reactogenicity was associated with anti-RBD seroconversion (p<0.001) but not anti-spike seroconversion. Two patients contracted COVID-19 following two doses of vaccine despite both having an anti-RBD antibody response above convalescent serum levels prior to anti-NP seroconversion. Both patients were hospitalized, but did not experience severe disease.

## Discussion

This prospective serology study found that while high rates of seroconversion were observed, consistent with other studies in hemodialysis patients,^12^ a robust anti-RBD response defined as reaching convalescent serum levels was seen in less than 10% of patients 28 days after a single dose. In contrast, two weeks after the second dose, 60% of hemodialysis patients had anti-RBD antibodies levels comparable to those achieved by infected patients. Anti-RBD response was lower than anti-spike response, which is of importance as anti-RBD may better correlate with viral neutralization.^9^ While our study did not evaluate cell-mediated immunity, a good anti-RBD response is required for adequate cell-mediated response.^13^ Interestingly, we found that symptoms following the second vaccine dose were associated with anti-RBD seroconversion and may help identify patients who develop some protection.

The response to the second dose, however, was notably weaker than in the HCW controls wherein 100% generated robust anti-RBD antibodies. This finding is similar to other compromised populations. In Canada, patients with malignancy and solid organ transplant receive two doses per manufacturer guidelines as studies demonstrating poor humoral response to one dose vaccination led to policy changes.^14,15^ With widespread global vaccine shortages, it is critical that we identify vulnerable populations. This study confirms a poor humoral response following a single dose of BNT162b2 COVID-19 vaccination in hemodialysis patients, and demonstrates the critical importance of the second dose, which should not be delayed in this vulnerable population.

## Data Availability

All data generated or analysed during this study are included in this published article (and its supplementary information files).

## Author Contributions

Dr. Hladunewich had full access to all of the data in the study and takes responsibility for the integrity of the data and the accuracy of the data analysis.

Concept and design: Yau, Perl, Bolotin, Tran, Gingras, Hladunewich.

Acquisition, analysis, or interpretation of data: All authors.

Drafting of the manuscript: Yau, Abe, Gingras, Hladunewich.

Critical revision of the manuscript for important intellectual content: All authors.

Statistical analysis: Naimark.

Administrative, technical, or material support: Mullin, Shadowitz, Quinn de Launay, Garnham-Takaoka, Takaoka, Colwill, Gingras.

Supervision: Hladunewich.

## Acknowledgements

We would like to thank Anny Gonzalez and Tatjana Sukovic for coordinating this study and the Sunnybrook Hemodialysis Staff for assisting with sample collection. We would also like to thank Shiva Barati, Darlene Cann, Gary Chao, Karen Green, Lois Gilbert, Nazrana Haq, Angel Xin Liu, Salma Sheikh Mohamed, Mohammad Mozafarihashjin, Aimee Paterson, Nimitha Paul, and Philip Samaan who assisted in data collection and management of samples from health care worker controls. We are grateful to Sharmistha Mishra, Stefan Baral, Adrienne Chan, Christine Fahim, Jennifer Gommerman, and Mario Ostrowski for sharing health worker data from the Wellness Hub initiative (#20-0339-E, Mount Sinai Hospital). The antibody measurements and initial data analysis were performed by Bhavisha Rathod, Mahya Fazel-Zarandi, Jenny Wang and Adrian Pasculescu. Convalescent serum data were obtained from the Toronto Invasive Bacterial Diseases Network (REB studies #20-044 Unity Health, #02-0118-U/05-0016-C, Mount Sinai Hospital).

