## Supplemental Tables and Figures for "The Humoral Response to the BNT162b2 Vaccine in Hemodialysis Patients"

### eTable 1. Comparison of Rates of SARS-CoV-2 IgG Anti-Spike and Anti-RBD Seroconversion in Hemodialysis Patients Receiving One Versus Two Doses of BNT162b2 and HCW Receiving Two Doses After Including and Excluding Individuals With Baseline Anti-NP Seroconversion

| Antibody | Study group | | Time point | Seroconversion No. (%)[a] | |
|---|---|---|---|---|---|
| | | | | Including baseline anti-NP seroconversion | Excluding baseline anti-NP seroconversion |
| Anti-spike | Hemodialysis | One dose | Pre-dose one | 8/66 (12%) | 6/63 (10%) |
| | | | Dose one + 4 weeks | 53/66 (80%) | 50/63 (79%) |
| | | Two dose[b] | Pre-dose two | 65/76 (86%) | 53/64 (83%) |
| | | | Dose two + 1 week | 72/76 (95%) | 60/64 (94%) |
| | | | Dose two + 2 weeks | 69/72 (96%) | 57/60 (95%) |
| | HCW | Two dose[b] | Dose two + 2-4 weeks | 35/35 (100%) | 35/35 (100%) |
| Anti-RBD | Hemodialysis | One dose | Pre-dose one | 2/66 (3%) | 0/63 (0%) |
| | | | Dose one + 4 weeks | 36/66 (55%) | 33/63 (52%) |
| | | Two dose[b] | Pre-dose two | 31/76 (41%) | 22/64 (34%) |
| | | | Dose two + 1 week | 58/76 (76%) | 47/64 (73%) |
| | | | Dose two + 2 weeks | 63/72 (88%) | 51/60 (85%) |
| | HCW | Two dose[b] | Dose two + 2-4 weeks | 35/35 (100%) | 35/35 (100% |

Abbreviations: HCW, health care worker; NP, nucleocapsid protein; RBD, receptor binding domain.
[a]Seroconversion threshold represents a positive test and are 0.19, 0.186, 0.396 for anti-spike, anti-RDB, and anti-NP respectively.
[b]Second dose was administered 21 days following first dose.

### eTable 2. Rates of Attaining Convalescent Serum Levels for SARS-CoV-2 IgG Anti-Spike and Anti-RBD in Hemodialysis Patients Receiving One Versus Two Doses of BNT162b2 and HCW Receiving Two Doses After Including and Excluding Individuals With Baseline Anti-NP Seroconversion

| Antibody | Study group | | Time point | Convalescent serum levels No. (%)[a] | |
|---|---|---|---|---|---|
| | | | | **Including baseline anti-NP seroconversion** | **Excluding baseline anti-NP seroconversion** |
| Anti-spike | Hemodialysis | One dose | Pre-dose one | 2/66 (3%) | 0/63 (0%) |
| | | | Dose one + 4 weeks | 15/66 (23%) | 13/63 (21%) |
| | | Two dose[b] | Pre-dose two | 19/76 (25%) | 14/64 (22%) |
| | | | Dose two + 1 week | 43/76 (57%) | 34/64 (53%) |
| | | | Dose two + 2 weeks | 52/72 (72%) | 43/60 (72%) |
| | HCW | Two dose[b] | Dose two + 2-4 weeks | 35/35 (100%) | 35/35 (100%) |
| Anti-RBD | Hemodialysis | One dose | Pre-dose one | 1/66 (2%) | 0/63 (0%) |
| | | | Dose one + 4 weeks | 4/66 (6%) | 2/63 (3%) |
| | | Two dose[b] | Pre-dose two | 10/76 (13%) | 7/64 (11%) |
| | | | Dose two + 1 week | 31/76 (41%) | 25/64 (39%) |
| | | | Dose two + 2 weeks | 43/72 (60%) | 35/60 (58%) |
| | HCW | Two dose[b] | Dose two + 2-4 weeks | 35/35 (100%) | 35/35 (100%) |

Abbreviations: HCW, health care worker; NP, nucleocapsid protein; RBD, receptor binding domain.
[a]The median level of antigen in convalescent serum taken 21-115 days post-symptom onset is considered a robust antibody response and are 1.38,1.25, and 1.13 for anti-spike, anti-RBD, and anti-NP antibodies respectively.
[b]Second dose was administered 21 days following first dose.

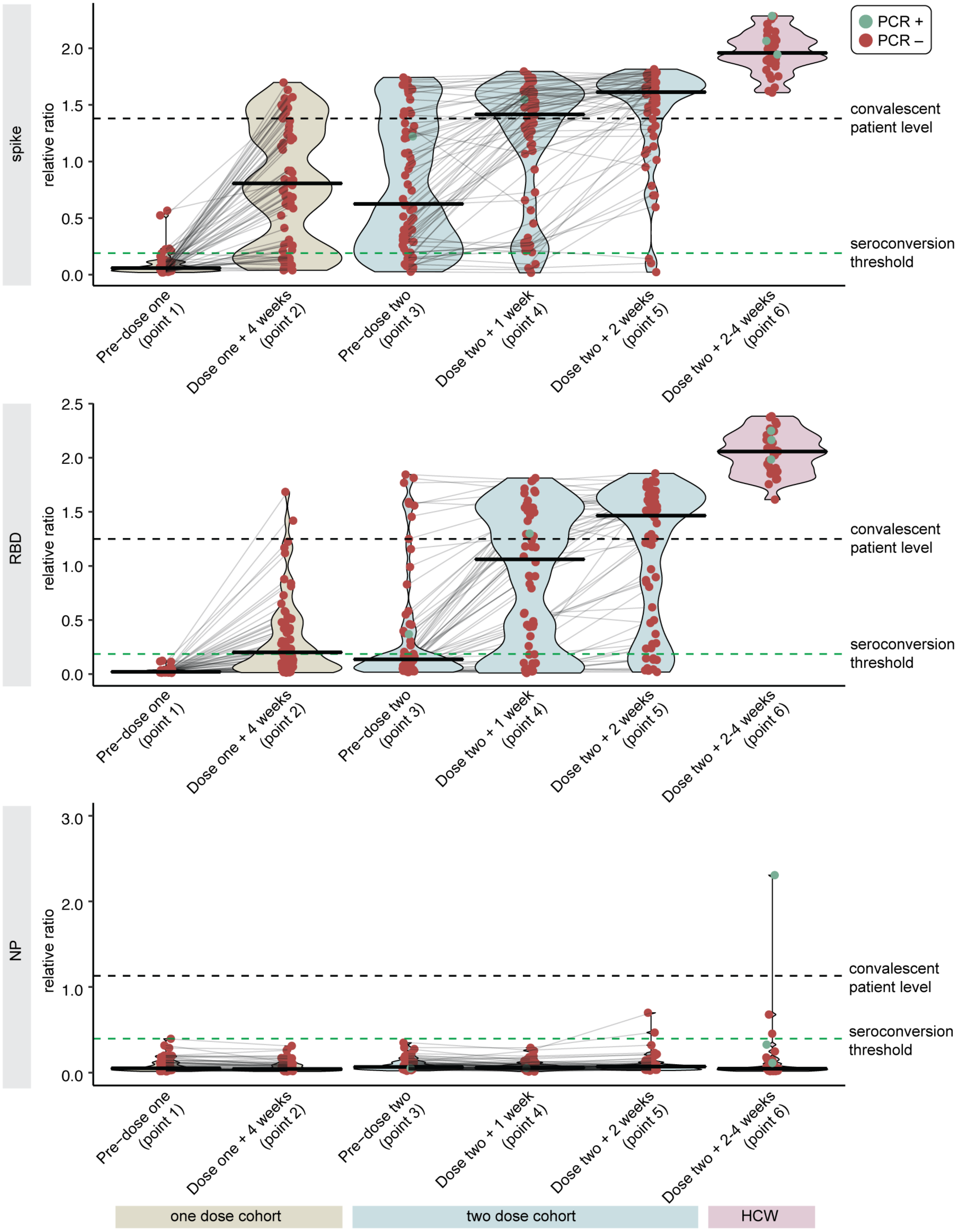
PCR +
PCR –
spike
RBD
NP
relative ratio
0.0
0.5
1.0
1.5
2.0
2.5
3.0
convalescent patient level
seroconversion threshold
Pre−dose one (point 1)
Dose one + 4 weeks (point 2)
Pre−dose two (point 3)
Dose two + 1 week (point 4)
Dose two + 2 weeks (point 5)
Dose two + 2-4 weeks (point 6)
one dose cohort
two dose cohort
HCW

### eFigure 1: SARS-CoV-2 IgG spike, RBD, and NP Antibody Response Following One Versus Two Dose BNT162b2 Vaccine in Hemodialysis Patients Excluding Baseline Anti-NP Seroconversion.

Antibody levels are reported as relative ratios to synthetic standards. The samples are grouped into two cohorts who received one (n=63) or two doses (n=64) of vaccine after excluding individuals with Anti-NP seroconversion at baseline. Dots represent individual serum samples collected at the indicated times, and the samples from the same patients are connected by lines. Green dots indicate individuals with prior RT-PCR confirmed SARS-CoV-2 infection. Seroconversion threshold represents a positive test and are 0.19, 0.186, and 0.396 for anti-spike, anti-RDB, and anti-NP antibodies respectively. The median level of antigen in convalescent serum taken 21-115 days post-symptom onset is considered a robust antibody response and is 1.38, 1.25, and 1.13 for anti-spike, anti-RBD, and anti-NP antibodies respectively. Healthy control workers (n=35) received two doses of vaccines with serologic measurement 2-4 weeks following dose 2. Abbreviations: HCW, health care worker; NP, nucleocapsid protein; RBD, receptor binding domain.
